# SARS-CoV-2 viral load distribution reveals that viral loads increase with age: a retrospective cross-sectional cohort study

**DOI:** 10.1101/2021.01.15.21249691

**Authors:** Sjoerd Euser, Sem Aronson, Irene Manders, Steven van Lelyveld, Bjorn Herpers, Jan Sinnige, Jayant Kalpoe, Claudia van Gemeren, Dominic Snijders, Ruud Jansen, Sophie Schuurmans Stekhoven, Marlies van Houten, Ivar Lede, James Cohen Stuart, Fred Slijkerman Megelink, Erik Kapteijns, Jeroen den Boer, Elisabeth Sanders, Alex Wagemakers, Dennis Souverein

**Affiliations:** Regional Public Health Laboratory Kennemerland, Haarlem, the Netherlands; Public Health Service Kennemerland, Haarlem, the Netherlands; Spaarne Gasthuis, Hoofddorp/Haarlem, the Netherlands; Comicro BV medical microbiology, Hoorn, the Netherlands; Department of Medical Microbiology, Noordwest Ziekenhuisgroep, Alkmaar, The Netherlands; Public Health Service Hollands Noorden, Alkmaar, the Netherlands; Rode Kruis Ziekenhuis, Beverwijk, the Netherlands; Wilhelmina Children’s Hospital, University Medical Center Utrecht, Utrecht, The Netherlands; Center for Infectious Disease Control, National Institute for Public Health and the Environment, Bilthoven, the Netherlands

**Author notes:** shared senior author. Corresponding author: Sjoerd M. Euser, Regional Public Health Laboratory Kennemerland, Boerhaavelaan 26, 2035 RC, Haarlem, the Netherlands, l.

**Keywords:** SARS-CoV-2, viral load distribution, age, COVID-19

## Abstract

**Objective:** To describe the SARS-CoV-2 viral load distribution in different patient groups and age categories.

**Methods:** All SARS-CoV-2 RT-PCR results from nasopharyngeal (NP) and oropharyngeal (OP) swabs (first PCR from unique patients only) that were collected between January 1 and December 1, 2020, predominantly in the Public Health Services regions Kennemerland and Hollands Noorden, province of North Holland, the Netherlands were included in this study. Swabs were derived from patients with respiratory symptoms who were presented at the general practitioner (GP), hospital, or hospital health care workers (HCWs) of four regional hospitals, nursing home residents and HCWs of multiple nursing homes, and in majority (>75%) from Public Health testing facilities of the two Public Health Services. SARS-CoV-2 PCR crossing point (Cp) values were used to estimate viral loads (higher Cp-values indicate lower viral loads).

**Results:** In total, 278.455 unique patients were tested of whom 9·1% (n=25.374) were SARS-CoV-2 positive. As there were differences in viral load distribution between tested populations, further analyses focused on PCRs performed by public health services (n=211.914) where sampling and inclusion were uniform. These data reveal a clear relation between age and SARS-CoV-2 viral load, with especially children aged<12 years showing lower viral loads than shown in adults (β: −0·03, 95CI% −0·03 to −0·02, p<0·001), independent of sex and/or symptom duration. Interestingly, the median Cp-values between the oldest (>79 years) and youngest (<12 years) population differed by over 4 PCR cycles, suggesting approximately a 16-fold difference in viral load. In addition, the proportion of children aged <12 years with a low load (Cp-value >30) was significantly higher compared to the other patients (31·1% vs. 17·2%, p-value<0.001).

**Conclusion:** We observed that in patients tested by Public Health Services, SARS-CoV2 viral load increases significantly with age. Previous studies suggest that young children (<12 years) play a limited role in SARS-CoV-2 transmission. Currently, the relation between viral load and infectivity is not yet well understood, and further studies should elucidate whether the lower viral load in children is indeed related to their suggested limited role in SARS-CoV-2 transmission. Moreover, as rapid antigen tests are less sensitive than PCR, these results suggest that SARS-CoV-2 antigen tests could have lower sensitivity in children than in adults.

## Introduction

Since the start of the COVID-19 pandemic, molecular testing of respiratory samples by reverse transcriptase polymerase chain reaction (RT-PCR) has been the primary method to diagnose SARS-CoV-2.^1^ Although SARS-CoV-2 RT-PCR results are in general reported in a qualitative manner (positive or negative), the quantitative test result (cycle threshold (Ct), or crossing point (Cp) which indicates the viral load in a sample) offers additional insights in for instance SARS-CoV-2 transmission dynamics. Individual viral load kinetics show a sharp increase in viral load in the earliest (mostly pre-symptomatic) stages of the infection, followed by a gradual decline.^2^ Interestingly, while viral cultures are mostly positive in samples with high viral load (Ct-value <25), samples with a low viral load hardly show any potential for viral cultivation (<3% at Ct 35), suggesting lower risk of transmission.^3^ One of the problems of studying SARS-CoV-2 viral loads in respiratory samples is the lack of comparability of Ct- or Cp-values derived from different laboratories, as these are assay- and method specific.^4^ This issue complicates the evaluation of SARS-CoV-2 viral loads in respiratory samples derived from large patient populations where often multiple laboratories are involved in analyzing these samples.

In this report we describe the SARS-CoV-2 viral load distribution of all routinely collected SARS-CoV-2 positive respiratory samples from a single large regional laboratory in the Netherlands, enabling us to evaluate the distinction between different patient groups (hospitalized patients, GP patients, nursing home patients, health care workers, patients tested in Public Health testing facilities) and age categories with respect to viral load distribution and the duration of symptoms.

## METHODS

### Setting, study design and participants

The Regional Public Health Laboratory Kennemerland, Haarlem, the Netherlands, performs SARS-CoV-2 RT-PCR testing for over 800.000 inhabitants, including health care workers (HCW), patients of four large teaching hospitals, patients of more than 600 GPs, 90 nursing home organizations, and those who are tested because of mild symptoms in public health testing facilities set up by the Public Health Services Kennemerland (PHS Kennemerland) and Hollands Noorden (PHS Hollands Noorden). Here, we report the SARS-CoV-2 RT-PCR results from nasopharyngeal (NP), oropharyngeal (OP) and combined swabs (first samples from unique patients only) that were analyzed between January 1 and December 1, 2020, using the RT-PCR based on the presence of the E-gene.^1^ Cp-values were calculated on Lightcycler 480 1.5.1 software (Roche diagnostics, Basel, Switzerland). Swabs were derived from GP patients, hospital patients and hospital health care workers (HCW), nursing home residents and nursing home HCWs, and in majority from Public Health testing facilities. As Public Health testing facilities employed a uniform sampling (combined oro- and nasopharynx) and inclusion policy, these PCRs were included for further analyses. Symptoms were required for Public Health testing in all test facilities, however between 13 August and 13 September, returning travelers without a need for symptoms were also tested at Amsterdam Airport Schiphol. For several months during the testing period, the national testing policy required children to also have severe symptoms (dyspnea or fever) or a positive contact to be included for testing (Supplemental text).

The Public Health Service Kennemerland, which is responsible for the data collection and reporting of new COVID-19 cases on a national level and performs contact tracing for these cases in the region Kennemerland, provided the date of first onset of disease for a subset of patients living in the region Kennemerland.

### Ethics Statement

The Medical Ethical Committee of the Amsterdam UMC approved this study on January 19^th^, 2021 (Study number: 2021.0022). In addition, the Institutional Review Board of the Spaarne Gasthuis, Hoofddorp/Haarlem, the Netherlands, approved to conduct the study on November 10th, 2020 (Study number: 2020.0154). The data were anonymized after collection and analyzed under code.

### Statistical analysis

Descriptive statistics were used to present the data: continuous variables were presented as median (interquartile range (IQR)), categorical variables were presented as no. (%). Comparisons of continuous variables were made using Mann-Whitney U tests, for categorical variables Chi-square tests were used. The Kruskal-Wallis test, combined with post-hoc pairwise comparisons with Bonferroni correction for multiple testing was used to investigate differences in viral load between different patientSor age groups. Linear regression analyses were used to analyze the relation between age and viral load allowing additional adjustment for potentially confounding variables. Statistical analyses were performed with R and RStudio (R version 4.0.3), packages tidyverse, sf and broom. P-values <0.05 were considered significant.

## RESULTS

Between January 1 and December 1, 278.455 unique patients were tested of whom the results of the first PCR were included. When patients were tested more than once, only the first positive PCR (when available) or the first negative PCR were included. Overall, 9·1% (n=25.374) samples were SARS-CoV-2 positive. Figure 1 shows the geographical distribution of the total number of positive unique patients for the period January 1-December 1, 2020 in the region Kennemerland and adjacent areas.

**Figure 1.**
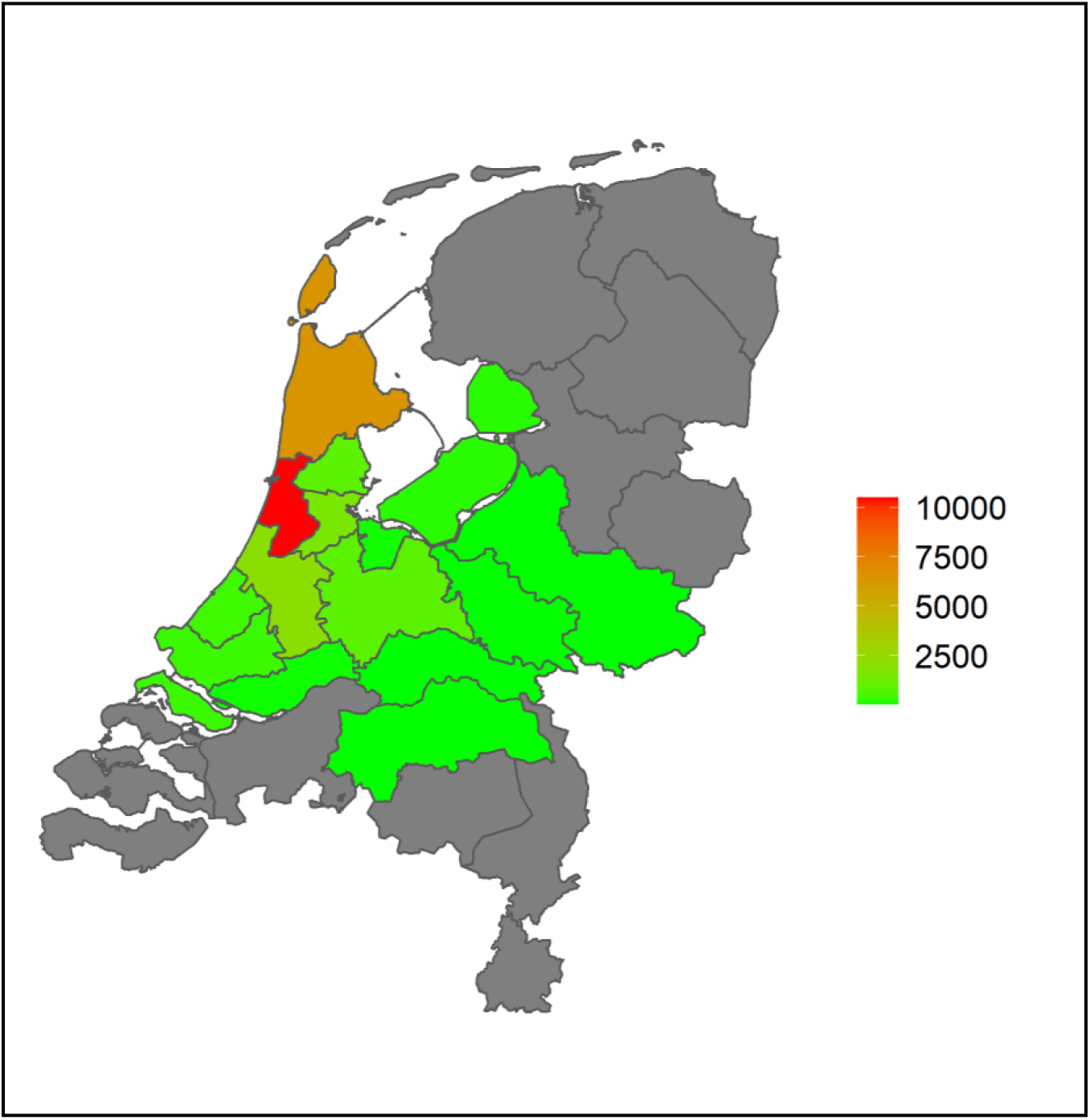
Geographical distribution of total number of SARS-CoV-2 PCR positive unique patients (n=25.374) tested in the Regional Public Health Laboratory Kennemerland for the period January-December 2020. Data are presented for Public Health Services areas; darker colors represent higher numbers of patients. The majority of patients lived relatively close by the laboratory, located in the red colored area.

### Viral load distribution in different patient populations

Comparison of the number of patients tested with the SARS-CoV-2 RT-PCR between the first (January 1-July 31) and second (August 1-December 1) wave of COVID-19 patients, reveals a shift in the number of tested patients from different patient populations (Table 1). In the first wave, a relatively large proportion of tests (26·6%) was performed for hospital patients, as compared to 0·8% in the second wave. The vast majority of samples (80·9%) in the second wave were derived from Public Health testing facilities.

**Table 1.**
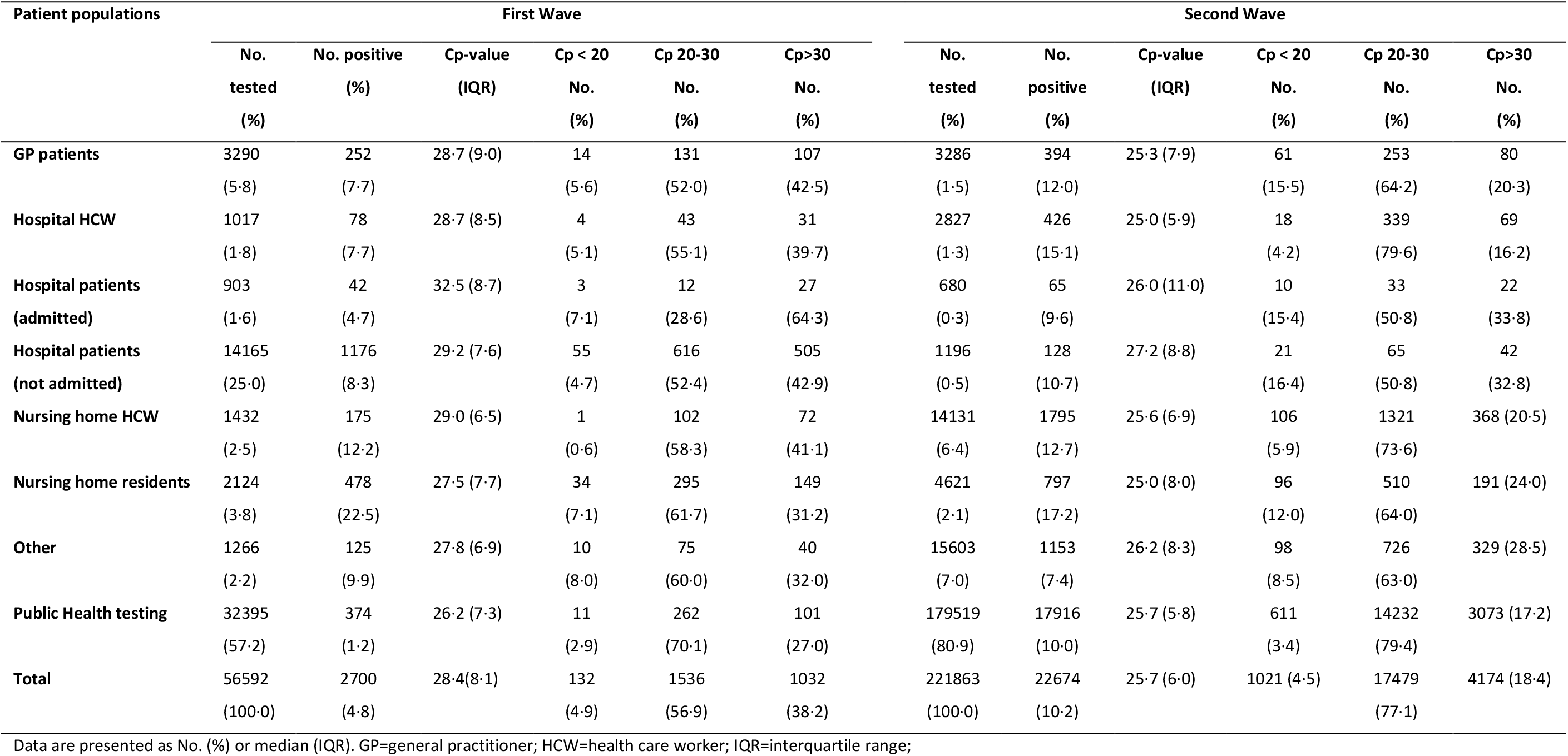
Cp-value characteristics for different patient populations in the First (January-July) and Second (August-December) Wave.

| Patient populations | First Wave |  |  |  |  |  | Second Wave |  |  |  |  |  |
| --- | --- | --- | --- | --- | --- | --- | --- | --- | --- | --- | --- | --- |
|  | No. | No. positive | Cp-value | Cp < 20 | Cp 20-30 | Cp>30 | No. | No. | Cp-value | Cp < 20 | Cp 20-30 | Cp>30 |
|  | tested<br>(%) | (%) | (IQR) | No.<br>(%) | No.<br>(%) | No.<br>(%) | tested<br>(%) | positive<br>(%) | (IQR) | No.<br>(%) | No.<br>(%) | No.<br>(%) |
| **GP patients** | 3290 | 252 | 28.7 (9.0) | 14 | 131 | 107 | 3286 | 394 | 25.3 (7.9) | 61 | 253 | 80 |
|  | (5.8) | (7.7) |  | (5.6) | (52.0) | (42.5) | (1.5) | (12.0) |  | (15.5) | (64.2) | (20.3) |
| **Hospital HCW** | 1017 | 78 | 28.7 (8.5) | 4 | 43 | 31 | 2827 | 426 | 25.0 (5.9) | 18 | 339 | 69 |
|  | (1.8) | (7.7) |  | (5.1) | (55.1) | (39.7) | (1.3) | (15.1) |  | (4.2) | (79.6) | (16.2) |
| **Hospital patients**<br>**(admitted)** | 903 | 42 | 32.5 (8.7) | 3 | 12 | 27 | 680 | 65 | 26.0 (11.0) | 10 | 33 | 22 |
|  | (1.6) | (4.7) |  | (7.1) | (28.6) | (64.3) | (0.3) | (9.6) |  | (15.4) | (50.8) | (33.8) |
| **Hospital patients**<br>**(not admitted)** | 14165 | 1176 | 29.2 (7.6) | 55 | 616 | 505 | 1196 | 128 | 27.2 (8.8) | 21 | 65 | 42 |
|  | (25.0) | (8.3) |  | (4.7) | (52.4) | (42.9) | (0.5) | (10.7) |  | (16.4) | (50.8) | (32.8) |
| **Nursing home HCW** | 1432 | 175 | 29.0 (6.5) | 1 | 102 | 72 | 14131 | 1795 | 25.6 (6.9) | 106 | 1321 | 368 (20.5) |
|  | (2.5) | (12.2) |  | (0.6) | (58.3) | (41.1) | (6.4) | (12.7) |  | (5.9) | (73.6) |  |
| **Nursing home residents** | 2124 | 478 | 27.5 (7.7) | 34 | 295 | 149 | 4621 | 797 | 25.0 (8.0) | 96 | 510 | 191 (24.0) |
|  | (3.8) | (22.5) |  | (7.1) | (61.7) | (31.2) | (2.1) | (17.2) |  | (12.0) | (64.0) |  |
| **Other** | 1266 | 125 | 27.8 (6.9) | 10 | 75 | 40 | 15603 | 1153 | 26.2 (8.3) | 98 | 726 | 329 (28.5) |
|  | (2.2) | (9.9) |  | (8.0) | (60.0) | (32.0) | (7.0) | (7.4) |  | (8.5) | (63.0) |  |
| **Public Health testing** | 32395 | 374 | 26.2 (7.3) | 11 | 262 | 101 | 179519 | 17916 | 25.7 (5.8) | 611 | 14232 | 3073 (17.2) |
|  | (57.2) | (1.2) |  | (2.9) | (70.1) | (27.0) | (80.9) | (10.0) |  | (3.4) | (79.4) |  |
| **Total** | 56592 | 2700 | 28.4(8.1) | 132 | 1536 | 1032 | 221863 | 22674 | 25.7 (6.0) | 1021 (4.5) | 17479 | 4174 (18.4) |
|  | (100.0) | (4.8) |  | (4.9) | (56.9) | (38.2) | (100.0) | (10.2) |  |  | (77.1) |  |

Data are presented as No. (%) or median (IQR). GP=general practitioner; HCW=health care worker; IQR=interquartile range;

In addition, the distribution of Cp-values (with higher Cp-values indicating a lower viral load) of respiratory samples showed lower viral loads for samples collected during the first wave compared to samples collected during the second wave (median Cp-value (IQR): 28·4 (8·1) vs. 25·7 (6·0), Mann-Whitney U test p-value<0·001) (Table 1). There were also remarkable differences in Cp-value distributions between patient populations who were tested in the first wave (Figure 2A) and the second wave of COVID-19 patients (Figure 2B). Analyses (Kruskal-Wallis) indicated significant differences in viral load between these patients populations in both the first (p-value<0·001) and second wave (p-value=0·001). Pairwise post-hoc comparisons (with a Bonferroni corrected p-value) showed significantly higher viral loads in the following patient populations in the first wave: Public Health (higher viral load) vs. GP patients (lower viral load) (p-value=0·022), Public Health vs. admitted hospital patients (p-value=0·001), Public Health vs. not admitted hospital patients (p-value<0·001), Public Health vs. nursing home HCWs (p-value<0·001), nursing home residents vs. not admitted hospital patients (p-value<0.001), nursing home residents vs. admitted hospital patients (p-value=0·007), nursing home residents vs. nursing home HCWs (p-value=0·016). In the second wave, nursing home residents vs. other groups (p-value=0·004), and GP patients vs. other groups (p-value=0·008) showed significantly higher viral loads (Supplemental table 1 and 2).

**Figure 2.**
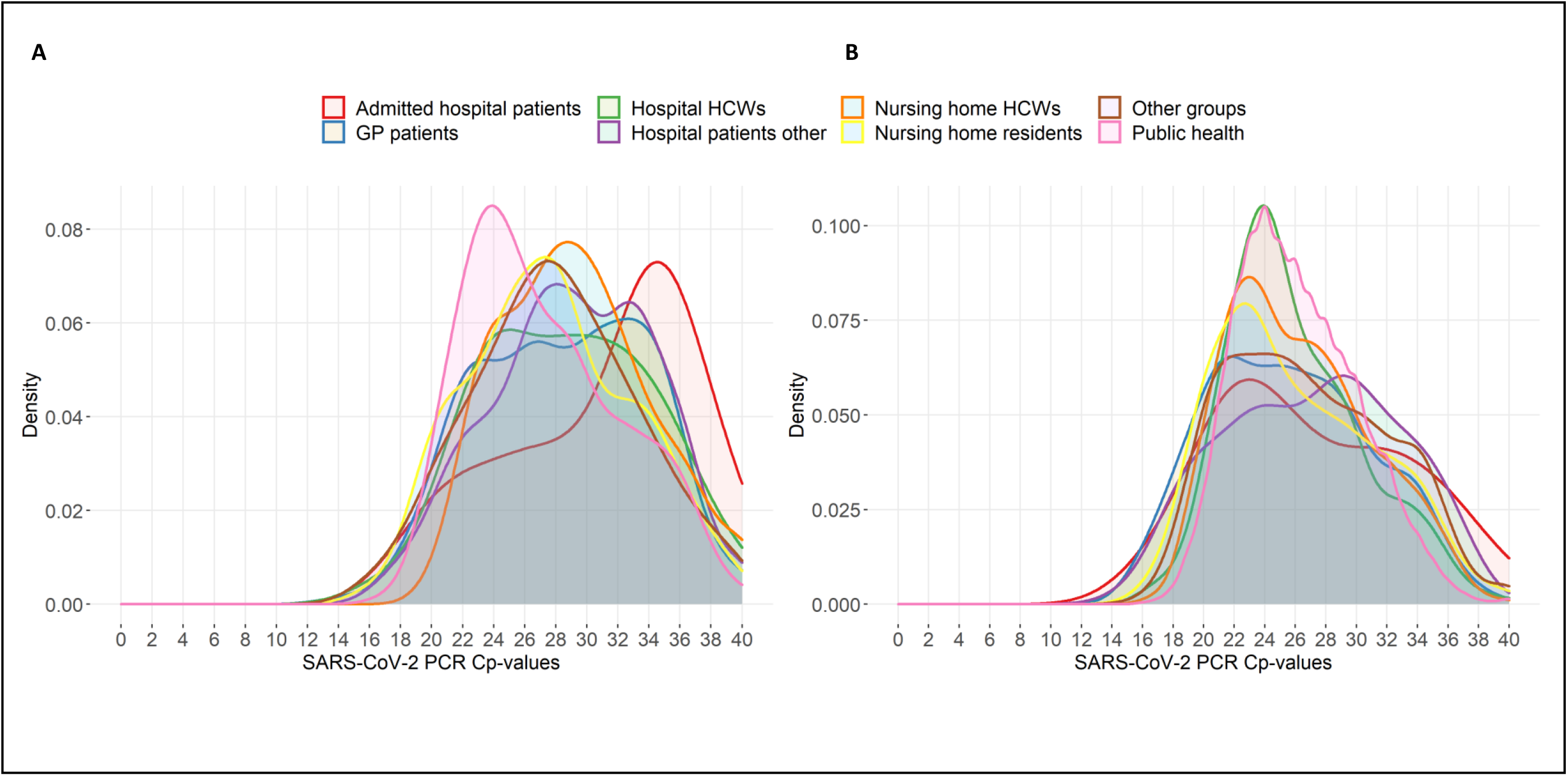
Distribution of SARS-CoV-2 PCR Cp-values within different patient populations in the first (Panel A, n=2.700) and second (Panel B, n=22.674) wave of the COVID-19 epidemic in the region Kennemerland and adjacent areas. Each color corresponds to one specific patient population that was routinely tested in the period January 1-July 31 (Panel A) or August 1-December 1 (Panel B). For each group the frequency of reported Cp-values was used to calculate a density score of which the area under the curve sums to 1. Higher Cp-values indicate lower viral loads, as they describe the number of PCR cycles at which the amplification signal crosses the fluorescence threshold.

### Viral load distribution across age and gender categories

The relation between patient age and SARS-CoV-2 viral load was investigated in the Public Health testing facilities samples collected between January 1 and December 1 (n=211.914), which was considered to be a relatively consistent population with respect to performed sampling procedure (a combined NP/OP swab by similarly trained professionals) and patient characteristics. In total, 8·6% (n=18.290) samples were SARS-CoV-2 positive. Patients were categorized in different age categories: age<12 years (primary grade school); 12-17 years (high school); 18-29 years; 30-49 years; 50-59 years; 60-69 years; 70-79 years; age>79 years. Number of available tests in each age category and proportion of positive tests are shown in Table 2. Distribution of the SARS-CoV-2 Cp-values for these age categories is shown in Table 2 and in Figure 3. These data showed that both proportion of positive tests as well as SARS-CoV-2 viral load increase with age. Proportion positive tests was 4·3% in children <12 years compared to 8·7% in all other patients (Chi-square, p-value<0·001). Analyses with a Kruskal-Wallis test indicated significant differences in viral load between the age categories (p-value<0·001). Pairwise post-hoc comparisons (with a Bonferroni corrected p-value) showed significant differences between almost all age categories, except for those within higher age groups (Supplemental table 3). In addition, the proportion of patients with a Cp-value>30 was highest in the younger age groups. For example, the proportion of children aged <12 years with a Cp-value >30 was significantly higher compared to the other patients (31·1% vs. 17·2%, Chi-square p-value<0·001). Interestingly, the median Cp-values had over 4 Cp-values difference between the oldest (>79 years) and youngest (<12 years) population, suggesting approximately 16 times difference in viral load. Additional data on the viral load distribution in age groups over time (March-December) are presented in Supplemental table 4. These showed that the relation between age and viral load was seen in all months, with a significant relation in months August-December (when the majority of tests was performed). To evaluate the differences between patients from the different Public Health Services, additional analyses on the relation between age groups and viral load were performed for each Public Health Service separately (Supplemental table 5) as well as for the returning travelers between 13 August and 13 September, who were tested at Amsterdam Airport Schiphol (n=6.033 of whom 74 (1·2%) travelers tested positive. These analyses showed that the relation between age and viral load distribution was independent of Public Health Service location.

**Table 2.** Cp-value characteristics for different age groups in Public Health patients.

| Age groups | Cp-values |  |  |  |  |  |
| --- | --- | --- | --- | --- | --- | --- |
|  | No. tested (%) | No. positive (%) | Cp-value<br>(IQR) | Cp < 20<br>No. (%) | Cp 20-30<br>No. (%) | Cp>30<br>No. (%) |
| <12 year | 5506 (2·6) | 238 (4·3) | 28·7 (5·0) | 0 (0) | 164 (68·9) | 74 (31·1) |
| 12-17 year | 22344 (10·5) | 1589 (7·1) | 26·9 (5·7) | 33 (2·1) | 1181 (74·3) | 375 (23·6) |
| 18-29 year | 47255 (22·3) | 4372 (9·3) | 26·0 (5·6) | 132 (3·0) | 3424 (78·3) | 816 (18·7) |
| 30-39 year | 41533 (19·6) | 2771 (6·7) | 25·9 (5·7) | 61 (2·2) | 2217 (80·0) | 493 (17·8) |
| 40-49 year | 34958 (16·5) | 3068 (8·8) | 25·5 (5·6) | 92 (3·0) | 2487 (81·1) | 489 (15·9) |
| 50-59 year | 30095 (14·2) | 3466 (11·5) | 25·0 (5·3) | 165 (4·8) | 2829 (81·6) | 472 (13·6) |
| 60-69 year | 19225 (9·1) | 1751 (9·1) | 25·3 (5·8) | 78 (4·5) | 1363 (77·8) | 310 (17·7) |
| 70-79 year | 8716 (4·1) | 800 (9·2) | 24·8 (5·9) | 48 (6·0) | 634 (79·2) | 118 (14·8) |
| >79 year | 2282 (1·1) | 235 (10·3) | 24·6 (5·4) | 13 (5·5) | 195 (83·0) | 27 (11·5) |
| **Total** | **211914 (100·0)** | **18290 (8·6)** | **25·7 (5·8)** | **622 (3·4)** | **14494 (79·2)** | **3174 (17·4)** |

Data are presented as No. (%) or median. IQR=interquartile range.

**Figure 3.**
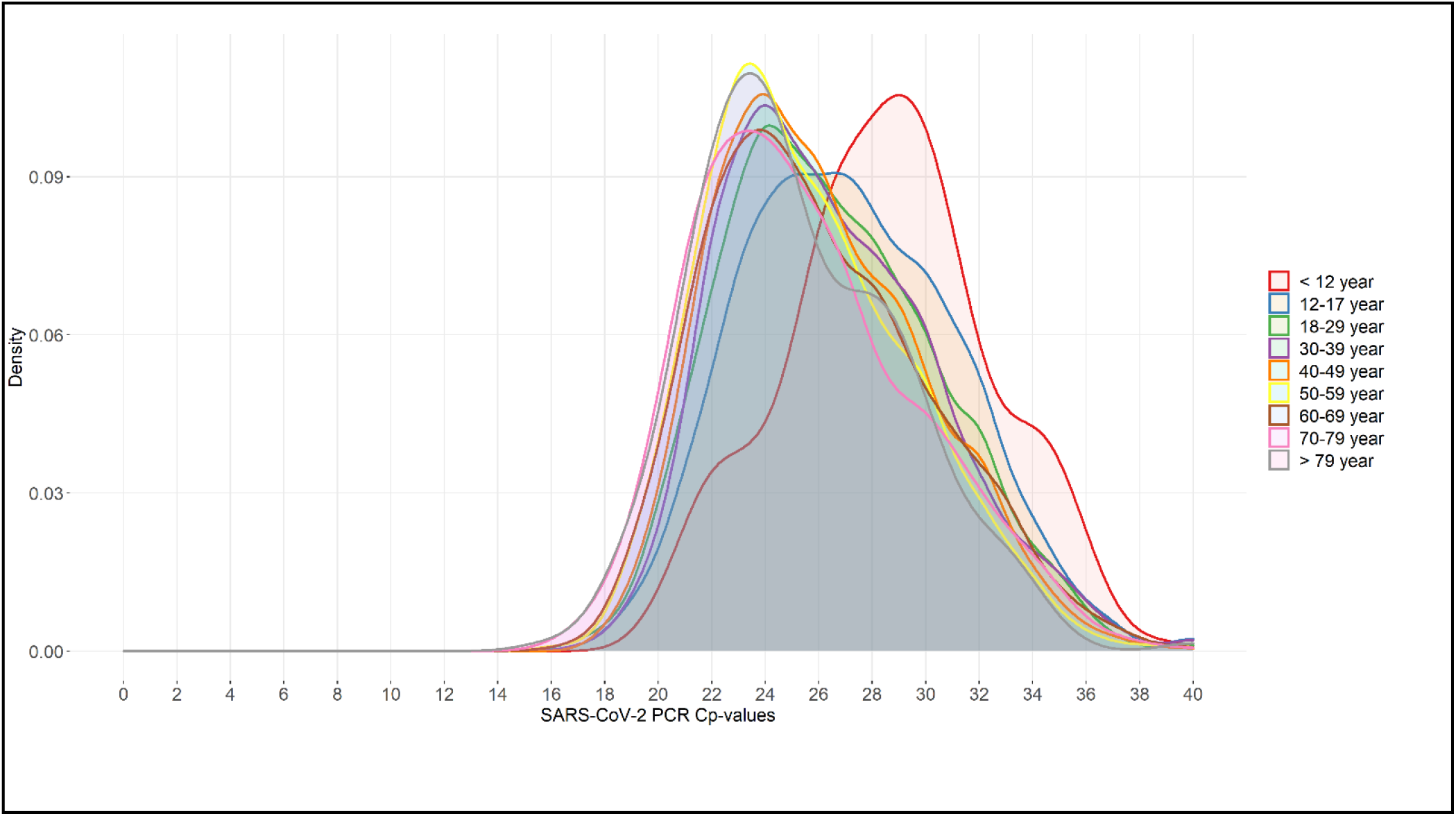
Distribution of SARS-CoV-2 PCR Cp-values within different age groups (n=18.290) of patients tested in a Public Health setting. Each color corresponds to one specific age group that was routinely tested in the period January 1-December 1. For each group the frequency of reported Cp-values was used to calculate a density score of which the area under the curve sums to 1.

When the non-Public Health Service SARS-CoV-2 positive samples (derived from all other patient populations) were analyses separately (n=6.931), there was a significant relation between age and viral load present in the OP swabs (linear regression, β: −0.01, 95CI%: −0·02 to 0·00, p-value<0·002) but not in the NP swabs (β: 0.00, 95CI%: −0·02 to 0·00, p-value=0·168) (Supplemental table 6).

In a subset of 7.300 samples from SARS-CoV-2 PCR positive Public Health patients (mean(SD) age: 40·7(17·8) years, 48·8% males) for whom time between onset of symptoms and testing was known (median (IQR) was: 2(3) days), linear regression analyses were performed. These analyses showed a significant relation between increasing age and decreasing Cp-values: β: −0·02, 95CI%: −0·03 to −0·02, p<0·001), independent of time between onset of symptoms and testing and sex (adjusted β: −0·03, 95CI%: −0·03 to −0·02, p<0·001). In addition, when the relation between patient sex and SARS-CoV-2 viral load was investigated (in the 7.300 SARS-CoV-2 PCR positive public health testing facilities samples), men showed a slightly (but significantly) higher viral load as compared to women (median Cp-value (IQR): 25·5 (5·9) vs. 25·8 (5·7), Mann-Whitney U test p-value=0·015), but when this was adjusted for age and time between onset of symptoms and testing in the linear regression analyses, no significant relation between sex and viral load was seen (adjusted β: 0·15, 95CI%: −0·03 to 0·33, p=0·096) anymore. Finally, a significant relation was found between viral load and time between onset of symptoms and testing (adjusted β: 0·45, 95CI%: 0·41 to 0·49, p<0·001), independent of age and sex.

## DISCUSSION

To our knowledge this is the first study to evaluate SARS-CoV-2 viral load distributions in a large number of patients from different patient categories. Our data present a clear relation between age and SARS-CoV-2 viral load, with children (<12 years) showing lower viral loads independent of sex and symptom duration. In addition, we observed that testing for COVID-19 using the SARS-CoV-2 RT-PCR in a large Dutch region shifted between the first wave (January 1-July 31) and second wave (August 1-December 1) of identified COVID-19 patients with respect to the populations of patients who were tested and the SARS-CoV-2 viral load distribution, with higher viral loads seen in samples analyzed in the second wave. The shift in tested patients from different patient populations between the first and the second wave was influenced by the national testing policy that was primarily focused on hospital patients in the first wave (due to limited testing capacity), and gradually moved to testing all patients with COVID-19 related symptoms (in large Public Health testing facilities) during the second wave. This could also have resulted in a more rapid diagnosis (since the onset of symptoms) for patients tested in the second wave resulting in the significantly higher viral load detected in the second wave samples.

Our observation of increasing SARS-CoV-2 viral load with increasing age, especially showing low viral loads in children under 12 years of age, is not in line with all previous studies.^5-8^ Previous studies were all smaller and suffered from heterogeneity in sampling technique, presence of symptoms, PCR technique (sometimes performed in different laboratories), symptom duration and test indication, and all included low numbers of pediatric patients. The largest of these, a cross-sectional study in Ghana including 9.549 positive samples from both symptomatic and asymptomatic patients at various health facilities, showed the lowest median viral loads in those aged ≤ 10 years (n=280) and the highest in those aged 71-80 years (n=110), without further statistical analysis.^9^ The second largest study (4.428 positive SARS-CoV-2 PCRs, including 96 patients born in or after 2000) did not show significant viral load differences in various age brackets, however the presence and duration of symptoms was unknown and the study comprised all sorts of materials (NP, OP, BAL, nasal wash etc.).^5^

The study of Jones et al.,^7^ who analyzed viral loads of 3.712 patients (of all ages) reported no significant differences in viral load across age groups, although their study population included only 117 patients aged<20 years, and it was discussed by others that there was in fact moderate evidence of increasing viral load with increasing age present in their data.^8^ In our study we had SARS-CoV-2 viral load data available for 18.290 unique patients tested in a public health setting, including 2.654 patients aged <20 years (with 238 children aged<12 years). A study in 1.213 positive specimens of symptomatic patients (with unknown symptom duration) only showed significantly lower Ct-values in the 80-89 years (n=86) age group.^10^ A study in 1.122 patients (type or setting unknown, tests performed in multiple laboratories) found a significantly higher age in high viral load samples (Ct-value<25: mean age 50), compared to moderate loads (Ct-value 25-30; mean age 48) and low viral loads (Ct-value >30; mean age 43), but when stratifying per age group the differences in high/moderate/low loads were not significant.^11^ Other studies were much smaller, showing various results and as at least as much heterogeneity compared to these larger studies.

There are several potential explanations for the lower SARS-CoV-2 viral load in respiratory samples derived from children. For instance, quality of swabbing might differ between children and adults. The discomfort caused by nasopharyngeal or oropharyngeal sampling in children could have resulted in more nasal or mid-turbinate samples, despite clear testing guidelines. There are studies that showed combined throat/nasal swab samples yielding a lower SARS-CoV-2 viral load compared to nasopharyngeal samples,^12^ although others showed no difference,^13^ or even higher viral loads in nasal or mid-turbinate samples.^14^ One could assume that even if the sampling method has a profound influence on SARS-CoV-2 viral load, this effect would have primarily been present in the youngest patients (where discomfort or protesting of the patient may have led to a different sampling procedure). As the effect of age on SARS-CoV-2 viral load in our data is present across a broad range of age-categories we do not think this can be explained by differences in sampling method alone. Moreover, to homogenize for sampling method, we only included PCRs performed by Public Health Services (combined OP/NP performed by trained personnel). By only including Public Health PCRs we also removed heterogeneity of inclusion criteria, although for children a restricted testing policy was employed during several months, where they needed to have severe symptoms (dyspnea/fever) or a positive contact. While it does explain the lower number of children tested, the increase in viral loads with age does not appear to change in time (Supplemental table 4), therefore changes in testing policy for children do not appear to explain our findings.

One could also argue that children were in general tested later after the first onset of symptoms compared to adults, as the parents postponed the moment of exposing their children to the distress of being sampled. This could have led to children being tested in a later stage of the infection, when their individual viral load may have declined compared to the early stage of the infections (following individual viral load kinetics).^2^ However, results were similar when we adjusted the relation between age and viral load for time since onset of disease.

Also, one could consider that despite the need for symptoms to be included for testing by public health agencies before December 1st, some of the tested population might have falsely reported symptoms in order to be tested free of charge. It is impossible to correct for this potential bias, and it is impossible to ascertain whether there are age-dependent differences in this respect. Nevertheless, it seems unlikely that the observed large difference in Cp-distribution is primarily caused by age-related differences in the false reporting of symptoms.

The observed lower viral load in children might be explained by age-related differences in viral infection dynamics. For example, several studies have suggested a differential expression of angiotensin-converting enzyme 2 (ACE2) (the receptor that SARS-CoV-2 uses for host entry) in different age categories.^15,16^ Bunyavanich et al. showed a positive association between ACE2 gene expression and age, which might explain the lower incidence of COVID-19 in children and the lower SARS-CoV-2 viral loads we found in the younger age categories.^15^ In addition, there are other factors that might protect children from higher viral load including for instance: differences in innate and adaptive immunity, more frequent recurrent and concurrent infections, pre-existing immunity to coronaviruses and differences in microbiota.^17^

Currently, antigen tests provide a rapid-yet less sensitive method to diagnose SARS-CoV-2. Antigen tests do not employ a target replication technique, and false-negatives are mostly observed in samples with a low viral load.^18^ We found Cp-values >30 in 31·1% of children <12 years, which was almost double the proportion found in the rest of the population. Therefore, these lower viral loads found in our study might indicate that antigen tests will have lower sensitivity in children. While further studies should validate these findings, caution should be warranted when using antigen tests to diagnose SARS-CoV-2 in populations with lower viral loads, in order to reduce the risk of false-negative results. One of the limitations of our study is that there were no data available on symptoms, underlying disease, sampling method, and moment of onset of first symptoms of all patients for whom respiratory samples were included. This is why the analyses of the relation between age and viral load was evaluated in samples from patients tested in the Public Health testing facilities which was considered to be a relatively consistent population with respect to performed sampling procedure and patient characteristics, and for whom time of onset of first symptoms was known (for a large subset of patients). Another limitation was the inclusion of symptomatic patients only, thus not reflecting the spectrum of SARS-CoV-2 infection, especially as asymptomatic presentations are frequently seen in children.^19–21^

In addition, it should be noted that the samples included in this study were collected before the novel SARS-CoV-2 variant, VOC 202012/01, that was first identified in the UK was likely to have been widespread in the Netherlands.^22^

To conclude, with this study we have tried to emphasize the usefulness of analyzing SARS-CoV-2 RT-PCR (viral load) data that are derived from a large population made up of a broad range of patient groups and age categories in a single laboratory (using the same SARS-CoV-2 RT-PCR method for all samples). With these data, shifts in tested patients populations and viral load distributions during the course of the COVID-19 pandemic can be closely monitored. This may contribute to a better understanding of SARS-CoV-2 transmission, and improve future measures that are taken to restrict viral spreading. The most remarkable finding of this study was the relation between SARS-CoV-2 viral load and age, with significantly lower viral loads in children. As previous studies have suggested that young children (<12 years) play a limited role in SARS-CoV-2 transmission,^8,21,23^ our data support this suggestion. Furthermore, these results suggest that SARS-CoV-2 antigen tests could have lower sensitivity in children than in adults.

However, viral load cannot solely explain differences in transmissibility between patients as for instance epidemiological aspects (exposure to others) and clinical presentation (coughing as a symptom) should not be overlooked.^6,8^ Further studies (combining viral load data with contact tracing data) should elucidate whether the lower viral load in children is indeed related to their suggested limited role in SARS-CoV-2 transmission.

## Data Availability

Data are available upon reasonable request

## Contributors

SE, SA, BH, AW and DS participated in conceptualisation. SE, IM, BH, JS, JK, RJ, IL, CG, DS, JCS, FSM, JB, AW and DS contributed to data collection. SE, AW and DS wrote the original draft. BH, SA, IM, SL, SSS, MH, EK, JB and ES contributed in reviewing and editing the paper. SE, IM and DS performed data curation and SE and DS contributed to data analysis. All authors critically reviewed and approved the final version.

## Acknowledgements

We would like to thank all physicians, nurses, Public Health testing personnel, laboratory technicians and administration personnel who have worked hard to provide SARS-CoV-2 testing for a large number of individuals, making it possible to perform these analyses.

## Supplementary material

### Supplemental text

#### Nationwide SARS-CoV2 PCR testing policy by Public Health authorities in The Netherlands

The most relevant changes in national testing policy in 2020 are summarized below.

#### 24-02-2020

One can be tested when they have Fever* (≥ 38 ° C, or feeling feverish in the elderly) AND at least one respiratory symptom (cough / dyspnoea) AND Symptoms arose within 14 days after (returning from an area with widespread transmission OR contact with a confirmed patient infected with SARS-CoV-2).

#### 06-03-2020

When family members of an index patient develop fever and/or respiratory symptoms they do not need to be tested, they will join in quarantine. Only contacts with a higher risk of severe disease (such as elderly and immunocompromised) need to be tested.

#### 12-03-2020

The need for epidemiological link to an area with widespread transmission or with a confirmed patient has been removed from the case definition. Outside hospitals, tests are only performed on persons with a higher risk of more severe disease when they develop severe disease, with fever AND respiratory symptoms (coughing or dyspnoea). These persons of higher risk include those ≥ 70 years old, those with co-morbidity (judged by indication for yearly influenza vaccination) and severely ill patients with respiratory symptoms and indication for hospital admission.

#### 19-03-2020

In (home) care institutions: after 1-2 proven cases of COVID-19, other patients are not tested. They need to be treated from the assumption that there is a COVID-19 outbreak in the institution.

#### 06-04-2020

Health care workers who provide direct patient care, with symptoms for ≥ 24 hours consistent with COVID-19 (coughing and/or fever and/ or rhinitis), can now be tested.

#### 06-05-2020

The need to have symptoms ≥ 24 hours to be tested is removed. A broader group of health care workers is now added to those eligible for testing. Children up to 12 years can be tested (upon approval of parents) when there are at least 3 children in their class/group with symptoms of COVID-19.

#### 01-06-2020

Everyone with symptoms of COVID-19 can be tested.

#### 13-08-2020 until 13-09-2020

Asymptomatic travellers from “code orange” countries / zones that land at Schiphol airport can be tested

#### 19-09-2020

Highest testing priority for persons with severe symptoms, followed by those from risk groups, followed by health care workers that provide care to risk groups, followed by symptomatic contacts of index patients. Teachers and health care workers will have priority testing. Testing asymptomatic persons has the lowest priority. Children under 12 with rhinitis can go to school, but have to stay in quarantine when they have a fever or dyspnoea. Children with symptoms of COVID-19 are only tested when they are severely ill, are contact of an index patient or part of an outbreak investigation.

#### 29-09-2020

Symptomatic children < 4 years do not need testing, unless they are severely ill or contacts of an index patient

#### 17-11-2020

Symptomatic children under 6 years are not by default tested. Children 6-12 years with only rhinitis can be tested, but it is not urgently adviced. Testing is urgently adviced when they are part of contact tracing, when they are severely ill (fever, dyspnoea or otherwise) or when testing is adviced due to an outbreak investigation.

#### 01-12-2020

(after the current study’s inclusion period)

Asymptomatic persons that are contacts of index patients can be tested at day 5 after the last contact.

**Supplemental table 1.** Kruskal-Wallis test post-hoc pairwise comparisons of Cp-values with Bonferroni correction between patient groups in the first wave (n=2.700)

|  | GP patients | Hospital<br>HCW | Hospital<br>patients<br>(admitted) | Hospital<br>patients<br>(not<br>admitted) | Nursing<br>home HCW | Nursing<br>home<br>residents | Other | Public<br>Health<br>testing |
| --- | --- | --- | --- | --- | --- | --- | --- | --- |
| GP patients | - | 1.000 | 0.342 | 1.000 | 1.000 | 0.744 | 1.000 | 0.022 |
| Hospital<br>HCW |  | - | 0.389 | 1.000 | 1.000 | 1.000 | 1.000 | 0.565 |
| Hospital<br>patients<br>(admitted) |  |  | - | 0.382 | 1.000 | 0.007 | 0.075 | 0.001 |
| Hospital<br>patients<br>(not<br>admitted) |  |  |  | - | 1.000 | <0.001 | 0.482 | <0.001 |
| Nursing<br>home HCW |  |  |  |  | - | 0.016 | 0.924 | <0.001 |
| Nursing<br>home<br>residents |  |  |  |  |  | - | 1.000 | 1.000 |
| Other |  |  |  |  |  |  | - | 1.000 |
| Public<br>Health<br>testing |  |  |  |  |  |  |  | - |

Data are p-values.

**Supplemental table 2.** Kruskal-Wallis test post-hoc pairwise comparisons of Cp-values with Bonferroni correction between patient groups in the second wave (n=22.674)

|  | GP patients | Hospital<br>HCW | Hospital<br>patients<br>(admitted) | Hospital<br>patients<br>(not<br>admitted) | Nursing<br>home HCW | Nursing<br>home<br>residents | Other | Public<br>Health<br>testing |
| --- | --- | --- | --- | --- | --- | --- | --- | --- |
| GP patients | - | 1.000 | 0.637 | 0.652 | 1.000 | 1.000 | 0.008 | 0.442 |
| Hospital<br>HCW |  | - | 1.000 | 1.000 | 1.000 | 1.000 | 0.120 | 1.000 |
| Hospital<br>patients<br>(admitted) |  |  | - | 1.000 | 1.000 | 1.000 | 1.000 | 1.000 |
| Hospital<br>patients<br>(not<br>admitted) |  |  |  | - | 1.000 | 1.000 | 1.000 | 1.000 |
| Nursing<br>home HCW |  |  |  |  | - | 1.000 | 0.195 | 1.000 |
| Nursing<br>home<br>residents |  |  |  |  |  | - | 0.004 | 0.469 |
| Other |  |  |  |  |  |  | - | 0.099 |
| Public<br>Health<br>testing |  |  |  |  |  |  |  | - |

Data are p-values.

**Supplemental table 3.** Kruskal-Wallis test post-hoc pairwise comparisons of Cp-values with Bonferroni correction between age groups (n=18.290)

|  | < 12 year | 12-17 year | 18-29 year | 30-39 year | 40-49 year | 50-59 year | 60-69 year | 70-79 year | >79 year |
| --- | --- | --- | --- | --- | --- | --- | --- | --- | --- |
| <12 year | - | <0.001 | <0.001 | <0.001 | <0.001 | <0.001 | <0.001 | <0.001 | <0.001 |
| 12-17 year |  | - | <0.001 | <0.001 | <0.001 | <0.001 | <0.001 | <0.001 | <0.001 |
| 18-29 year |  |  | - | 1.000 | <0.001 | <0.001 | <0.001 | <0.001 | <0.001 |
| 30-39 year |  |  |  | - | 0.020 | <0.001 | 0.010 | <0.001 | 0.001 |
| 40-49 year |  |  |  |  | - | <0.001 | 1.000 | 0.005 | 0.103 |
| 50-59 year |  |  |  |  |  | - | 0.059 | 1.000 | 1.000 |
| 60-69 year |  |  |  |  |  |  | - | 0.084 | 0.325 |
| 70-79 year |  |  |  |  |  |  |  | - | 1.000 |
| >79 year |  |  |  |  |  |  |  |  | - |

Data are p-values.

**Supplemental table 4.**
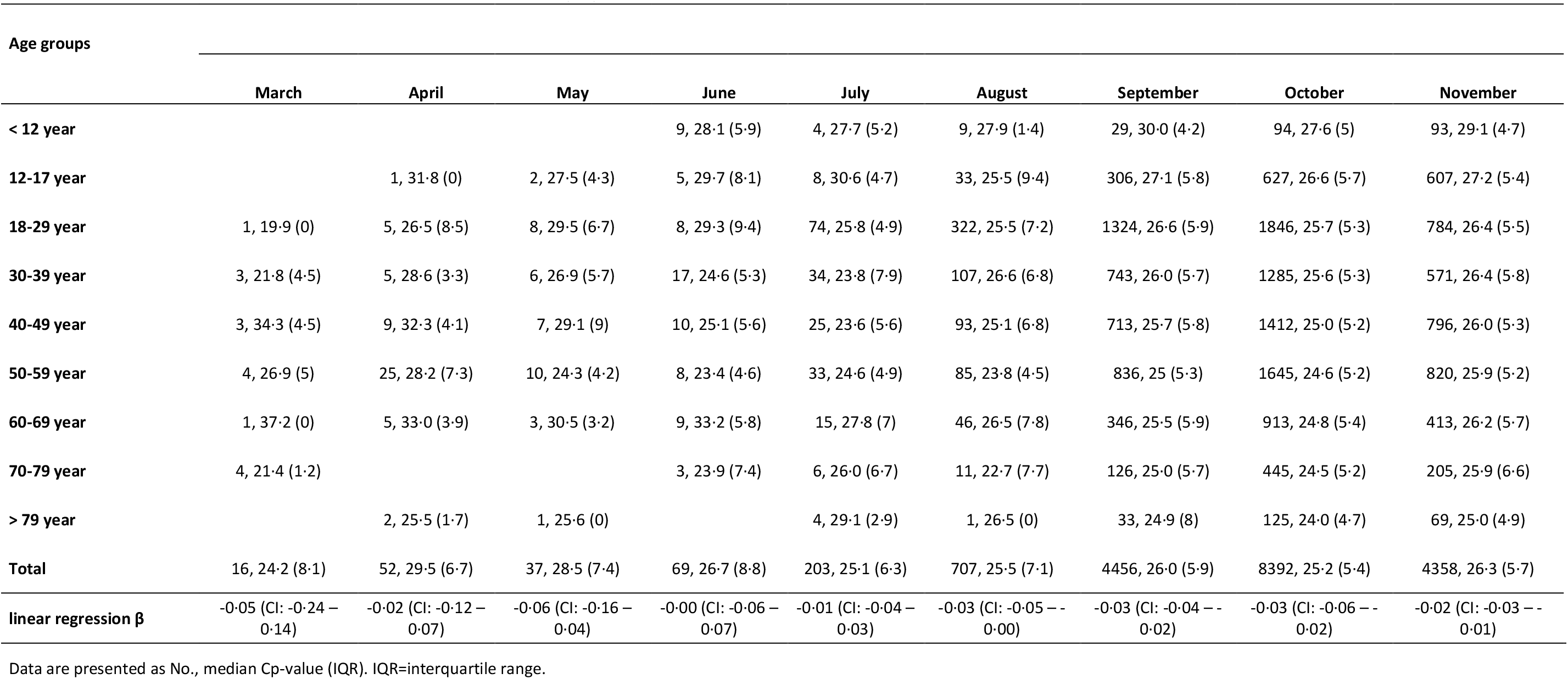
Cp-value characteristics for different age groups per month.

**Supplemental table 5.** Cp-value characteristics for different age groups and Public Health Services.

| Age groups |  |  |  |  |
| --- | --- | --- | --- | --- |
|  | PHS Hollands Noorden | PHS Kennemerland<br>airport arrivals | PHS Kennemerland without<br>airport arrivals | PHS Other |
| < 12 year | 78, 29·0 (4) | - | 146, 28·1 (5) | 14, 30·1 (5·6) |
| 12-17 year | 600, 27·3 (5·5) | - | 895, 26·7 (6) | 94, 26·8 (4·4) |
| 18-29 year | 1516, 26·1 (5·4) | 47, 26·0 (8·5) | 2540, 25·9 (5·8) | 269, 26·7 (5·7) |
| 30-39 year | 839, 26 (5·3) | 12, 27·5 (11·4) | 1796, 25·8 (5·8) | 124, 26·5 (6·9) |
| 40-49 year | 1050, 25·6 (5·5) | 2, 24·4 (3·5) | 1860, 25·3 (5·6) | 156, 26·1 (5·6) |
| 50-59 year | 1286, 24·9 (5·2) | 8, 26 (6·8) | 2015, 25·0 (5·4) | 157, 25·0 (6·9) |
| 60-69 year | 742, 25·6 (6·1) | 4, 22·1 (5·5) | 938, 25·2 (5·7) | 67, 26·0 (6) |
| 70-79 year | 350, 24·9 (5·5) | - | 424, 24·7 (6) | 26, 25·7 (5·9) |
| > 79 year | 87, 24·5 (4·6) | 1, 23·9 (0) | 138, 24·7 (5·6) | 9, 24·8 (8·3) |
| Total | 6548, 25·8 (5·6) | 74, 26·0 (8·9) | 10752, 25·6 (5·8) | 916, 26·3 (6·4) |
| linear regression $\beta$ | -0·03 (CI: -0·03 – -0·02) | -0·02 (CI: -0·11 – 0·07) | -0·02 (CI: -0·03 – -0·02) | -0·03 (CI: -0·04 – -0·01) |

Data are presented as No., median Cp-value (IQR). IQR=interquartile range.

**Supplemental table 6.** Cp-value characteristics for different age groups and sampling material within non-public Health patienta.

| Age groups | Nasopharyngeal (NP) | Oropharyngeal (OP) |
| --- | --- | --- |
| < 12 year | 9, 27.8 (11.8) | 23, 27.8 (6.5) |
| 12-17 year | 28, 26.8 (5.7) | 40, 28 (9.3) |
| 18-29 year | 499, 26 (6.7) | 439, 27.3 (8.1) |
| 30-39 year | 438, 25.3 (7.1) | 374, 26.8 (7.5) |
| 40-49 year | 484, 25.3 (7) | 404, 28 (8.7) |
| 50-59 year | 607, 24.8 (6.8) | 764, 27.9 (8) |
| 60-69 year | 300, 25.9 (7.7) | 481, 28 (8.3) |
| 70-79 year | 87, 26.2 (8.5) | 525, 27.7 (9.2) |
| > 79 year | 212, 25 (8.3) | 1247, 26.6 (8) |
| Total | 2664, 25.4 (7.2) | 4297, 27.3 (8.4) |
| linear regression $\beta$ | 0.00 (CI: -0.02 – 0.00)<br>p-value=0.168 | -0.01 (CI: -0.02 – -0.00)<br>p-value=0.002 |

Data are presented as No., median Cp-value (IQR). IQR=interquartile range.

**Supplemental Figure 1:**
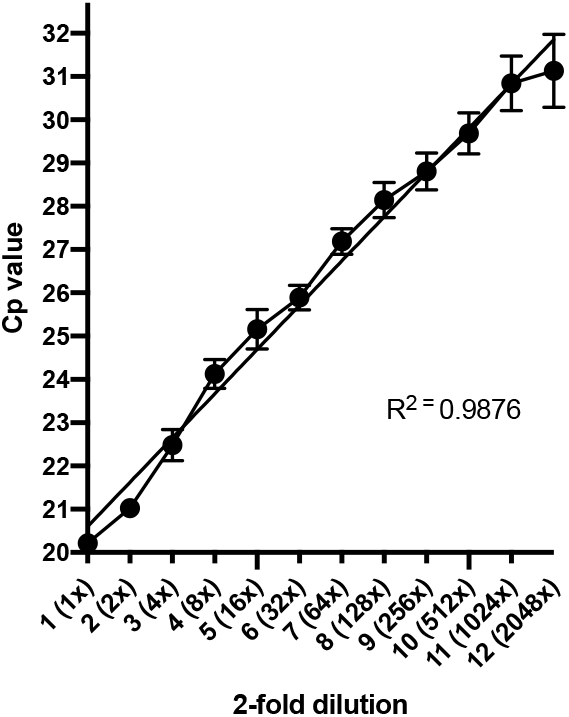
A 1:1 dilution series (across median Cp-value range in various age groups) was performed in threefold using a positive SARS-CoV-2 sample. Cp-value versus SARS-CoV-2 dilution correlation was analysed, with R-squared of 0.9876, with a slope of 1,023 Cp difference per 1:1 dilution.

## REFERENCES

1 Corman VM, Landt O, Kaiser M, et al. Detection of 2019 novel coronavirus (2019-nCoV) by real-time RT-PCR. Euro Surveill. 2020; 25(3): pii=2000045. https://doi.org/10.2807/1560-7917.ES.2020.25.3.2000045.

2 Wölfel R, Corman VM, Guggemos W, et al. 2020. Virological assessment of hospitalized patients with COVID-2019. Nature 2020; 581: 465–73. https://doi.org/10.1038/s41586-020-2196-x.

3 Jaafar R, Aherfi S, Wurtz N, et al. Correlation between 3790 qPCR positives samples and positive cell cultures including 1941 SARS-CoV-2 isolates. Clin Infect Dis 2020; https://doi.org/10.1093/cid/ciaa1491.

4 Rhoads D, Peaper DR, She RC, et al. College of American Pathologists (CAP) Microbiology Committee Perspective: Caution Must Be Used in Interpreting the Cycle Threshold (Ct) Value. Clin Infect Dis 2020; https://doi.org/10.1093/cid/ciaa1199.

5 Kleiboeker S, Cowden S, Grantham J, et al. SARS-CoV-2 viral load assessment in respiratory samples. J Clin Virol 2020;129: 104439; https://doi.org/10.1016/j.jcv.2020.104439.

6 Jacot D, Greub G, Jaton K, Opota O. Viral load of SARS-CoV-2 across patients and compared to other respiratory viruses. Microbes and Infection 2020; 22: 617–21.

7 Jones TC, Mühlemann B, Veith T, et al. An analysis of SARS-CoV-2 viral load by patient age. 2020; https://doi.org/10.1101/2020.06.08.20125484.

8 Walsh KA, Jordan K, Clyne B, et al. SARS-CoV-2 detection, viral load and infectivity over the course of an infection. J of Infection 2020; 81: 357–71.

9 Owusu M, Sylverken AA, Ankrah ST., et al. Epidemiological profile of SARS-CoV-2 among selected regions in Ghana: A cross-sectional retrospective study. PLOS ONE 2020; https://doi.org/10.1371/journal.pone.0243711.

10 Buchan B, Hoff JS, Gmehlin CG, et al. Distribution of SARS-CoV-2 PCR Cycle Threshold Values Provide Practical Insight Into Overall and Target-Specific Sensitivity Among Symptomatic Patients. Am J Clin Pathol 2020;154:479–85.

11 Maltezou HC, Raftopoulos V, Vorou R, et al. Association between upper respiratory tract viral load, comorbidities, disease severity and outcome of patients with SARSCoV-2 infection. J Infect Dis 2021; https://academic.oup.com/jid/advance-article/doi/10.1093/infdis/jiaa804/6060070.

12 Vlek ALM, Wesselius TS, Achterberg R, Thijsen. Combined throat/nasal swab sampling for SARS-CoV-2 is equivalent to nasopharyngeal sampling. Eur J Clin Microbial Infect Dis 2021; 40: 193–5. https://doi.org/10.1007/s10096-020-03972-y.

13 Péré H, Podglajen I, Wack M, et al. Nasal Swab Sampling for SARS-CoV-2: a Convenient Alternative in Times of Nasopharyngeal Swab Shortage. J Clin Microbiol 2020; 58(6): e00721–20; https://jcm.asm.org/content/58/6/e00721-20.

14 Tu YP, Jennings R, Hart B, et al. Swabs Collected by Patients or Health Care Workers for SARS-CoV-2 Testing. NEJM 2020; 383: 5; https://www.nejm.org/doi/full/10.1056/NEJMc2016321.

15 Bunyavanich S, Do A, Vicencio A. Nasal Gene Expression of Angiotensin-Converting Enzyme 2 in Children and Adults. JAMA 2020;323(23): 2427–29; https://jamanetwork.com/journals/jama/fullarticle/2766524.

16 Patel AB, Verma A. Nasal ACE2 levels and Covid-19 in children. JAMA 2020;323(23):2386–87; https://jamanetwork.com/journals/jama/fullarticle/2766522.

17 Zimmermann P, Curtis N. Why is COVID-19 less severe in children? A review of the proposed mechanisms underlying the age-related difference in severity of SARS-CoV-2 infections. Arch Dis Child 2020; doi:10.1136/archdischild-2020-320338.

18 Porte L, Legarraga P, Vollrath V, et al. Evaluation of a novel antigen-based rapid detection test for the diagnosis of SARS-CoV-2 in respiratory samples. Int J Infect Dis 2020; 99:328–33.

19 Baggio S, L’Huillier AG, Yerly S, et al. SARS-CoV-2 viral load in the upper respiratory tract of children and adults with early acute COVID-19. Clin Infect Dis 2020; doi:10.1093/cid/ciaa1157.

20 Zimmermann P, Curtis N. COVID-19 in Children, Pregnancy and Neonates: A Review of Epidemiologic and Clinical Features. Pediatr Infect Dis J 2020;39:469–77.

21 Williams PCM, Howard-Jones AR, Hsu P, et al. SARS-CoV-2 in children: spectrum of disease, transmission and immunopathological underpinnings. Pathology 2020; 52(7): 801–808.

22 Davies NG, Barnard RC, Jarvis CI, et al. Estimated transmissibility and severity of novel SARS-CoV-2 Variant of Concern 202012/01 in England. 2020; https://www.medrxiv.org/content/10.1101/2020.12.24.20248822v1.

23 Gudbjartsson DF, Helgason A, Jonsson H, et al. Spread of SARS-CoV-2 in the Icelandic population NEJM 2020;382:2302–15; https://www.nejm.org/doi/pdf/10.1056/NEJMoa2006100?articleTools=true.

